# Impaired Capillary Endothelial Cell Differentiation Contributes to pulmonary hypertension in a dynamic Capillary-Alveoli Micro-physiological System and animal models

**DOI:** 10.64898/2026.02.21.26346776

**Authors:** Yaning Li, Xiaomeng Liu, Pei Mao, Tengfei Zhou, Xiaofang Fan, Guoyu Xie, Yixin Ji, Weiwei Wang, Gang Han, Jun Jiang, Chao Zhang, Jun Yang

**Author notes:** Correspondence to: Jun Yang. These authors contributed equally: Yaning Li, Xiaomeng Liu.

## Abstract

Pulmonary hypertension (PH) is a progressive condition characterized by increased pulmonary arterial pressure. Endothelial cell dysfunction is one important characteristic of PH. Recently, capillary endothelial cells, including aerocytes (aCaps) and general capillary cell (gCaps), have been detected in developing lungs but their role and the regulatory mechanisms underlying PH remain poorly understood.

The goal of this study was to identify changes in Caps and their effects on hypertensive pulmonary circulation. We set up a Capillary Alveoli Micro-physiological System (CAMS) incorporated with hPSCs(human pluripotent stem cells)-aCaps to show loss of Cap connection under dynamically cultured hypoxic condition. We employed single-cell RNA sequencing (scRNA-seq) and immunofluorescence to demonstrate impaired gCaps differentiation with increased expression of cell membrane receptor CD93 in PH patients and a Sugen 5416/hypoxia (SuHx) rat model. Conditional Knockdown or Lentiviral overexpression of CD93 alleviated the pathology observed in SuHx mice. We also revealed that CD93 overexpression upregulated SMAD2/3 to repress Apelin (APLN) expression by CHIP assay. Finally, supplementation with an APLNR agonist in the PH rat model promoted gCaps-to-aCaps differentiation and improved haemodynamic indices.

Overall, our results highlight the potential for promoting capillary cell differentiation with G protein biased APLNR agonist as a therapeutic strategy for pulmonary vascular disease.

## Introduction

Capillary networks are the primary site for the delivery of oxygen, removal of metabolic waste, and the control of blood flow which has been implicated in a range of pathologies including pulmonary diseases^1^. Capillary vessels are composed of a single layer of endothelial cells (ECs) ^2^. Endothelial dysfunction and muscularized pulmonary microvasculature are important characteristics of pulmonary hypertension (PH), a progressive condition with mean pulmonary arterial pressure (mPAP) exceeding 20 mmHg that finally leads to right heart failure^3^. It can be caused by physiological (high-altitude) or pathological (respiratory) , and hypoxia ^4^. The precise control of blood flow through capillary beds, governed in part by dynamic capillary dilation, is critical for gas diffusion^5, 6^. Although the major pathological thickness of pulmonary arterial wall could be caused by anti-apoptotic ECs clones formation, or capillary cells migration^7^, the alteration at pulmonary capillary beds in PH hasn’t been clearly demonstrated.

Loss of capillaries has been reported in PH patients with low diffusing capacity of the lung for carbon monoxide (DLCO)^8^, ventilation□perfusion (V/Q ratio) mismatch may also play a role in first class of PH. Since capillary bed dilation has been frequently observed in genetic form of PH patients, the dilated Capillary bed probably has some impact on diffusion, which warrants further investigation^9^.

Recently, a new type of pulmonary capillary cell – aerocytes (aCaps) was identified with which intermixed general capillary cells (gCaps), which were positive for the transmembrane phosphoglycoprotein CD34^10^. gCaps are believed to have stem/progenitor-like properties that contribute to capillary stability and self-renewal^10^. Single-cell RNA sequencing (scRNA-seq) analysis of lung tissue from PH patients and a Sugen 5416-hypoxia (SuHx) rat model revealed increased proportion of gCaps ^11^. However, analysis of differentially expressed genes (DEGs) in the lung tissue of PH patients revealed significant downregulation of genes associated with stemness regulation (*KLF4*, *ATF3*, and *TCF4*) in gCaps^12–15^. These cells have never been successfully derived and cultured separately in vitro, which makes the mechanism study of Caps in PH challenging.

ACaps are specialized for gas exchange ^10^ and its involvement in PH only be suggested in mice so far ^11, 16^. Apelin (APLN), a secreted polypeptide, functions as a marker for aCaps. Although treatment with APLN ameliorated PH in clinical trials, the side effect on the heart delayed its approval^17–19^, and whether APLN acts through its receptor APLNR on gCaps and affects its function remains to be clarified.

The goal of this study was to elucidate the transition of Caps and the precise mechanisms responsible for the altered capillary bed in PH. Surprisingly, our results suggested a reduction in aCaps differentiation in the lungs of PH patients. We set up a Capillary-Alveoli Micro-physiological System (CAMS) incorporated with human pluripotent stem cells(hPSCs) differentiated aCaps to detect hypoxia related signal. With single cell RNA sequencing(scRNA-seq), we identified CD93 as a highly expressed cell membrane receptor in gCaps; therefore, we generated a lineage tracing mice to study the role of CD93 in pathogenesis. By combining lentiviral overexpression and CRISPR-Cas9 knockout, we demonstrated that CD93 regulated the activity of SMAD2/3-APLN singaling pathway. Unlike direct administration of APLN, administration of G-protein-biased APLNR agonist prevented and reversed PH in SuHx rats without impacting cardiac hypertrophy. Overall, our results demonstrate that a reduction in gCaps-to-aCaps differentiation occurred in the lungs of PH and loss of capillary network on dynamically cultured human organoids of CAMS under hypoxia condition, that APLNR agonist may ameliorate PH by promoting the differentiation of gCaps into aCaps.

## Results

### CD34^+^ Caps contribute to dilated pulmonary capillary beds and mediate the differentiation into aCaps from hPSCs

To investigate the role of capillary endothelial progenitor cells in pulmonary vascular disease and recapitulate their formation with human PSCs according the understanding of their generation, which hasn’t been documented yet^20^. We first conducted immunochemistry (IHC) on lung tissue samples from 3 PH patients and 3 controls. Staining revealed notably vascular remodeling and dilated capillary beds in the lungs of PH patients (Fig. 1, A; and Fig. S1, A and B). Further, we performed CD31-staining to identify endothelial cells(ECs), 3D imaging of CD31-stained and cleared tissues confirmed capillary dilation in alveolar regions (Fig. 1, B and C), consistent with the finding of a previous study^8^. Because capillary cells has no muscular layer at normal condition, whereas our costaining of CD34 with α-SMA or CD31 showed that CD34⁺ capillary cells were wrapped with smooth muscle cells in muscularized capillaries from PH patients, together with increased percentage of CD34^+^ CD31^+^cells and elevated *CD34* mRNA levels (Fig. 1, D-F; and Fig. S1, C), which was also observed in a Sugen 5416/hypoxia (SuHx) rat model (Fig. S2). It suggested that CD34⁺ cells contribute to vascular pathology by promoting the development of an EC progenitor phenotype and could also increase the number of capillary ECs. To verify these alveolar capillary alterations at the transcriptional level, we used quantitative real-time PCR (qRT-PCR) to quantify the expression of aCaps markers including *APLN, EDNRB, HPGD* and *CAR4*. We detected reduced expression of aCaps marker expression (Fig. 1, G), suggesting that the reduction in aCaps was accompanied by an increase in other CD34⁺ cells, such as capillary endothelial progenitor cells in altered alveolar-capillary in the alveoli of PH patients.

**Figure 1.**
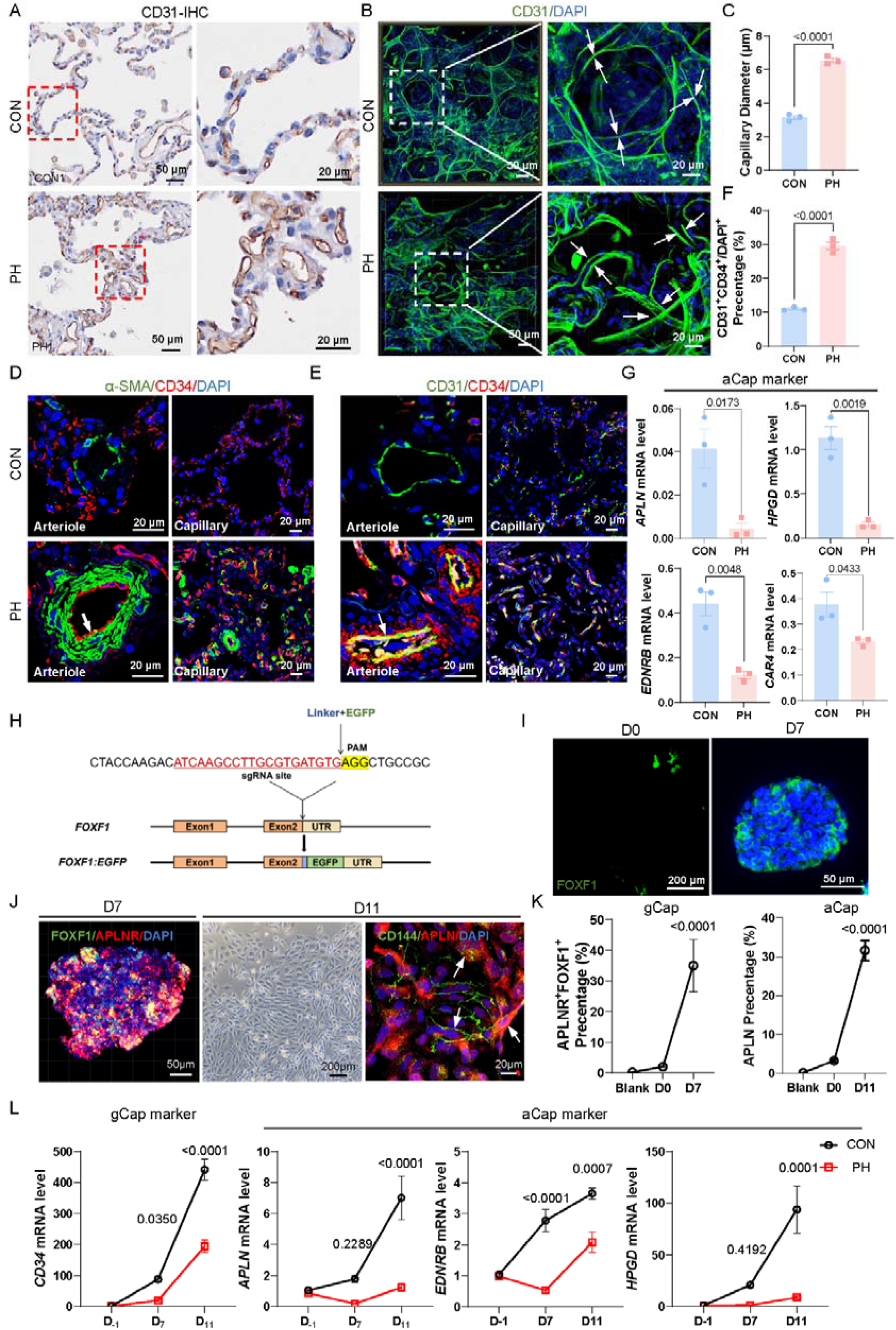
CD34^+^ Caps contribute to dilated pulmonary capillary beds and mediate the differentiation into aCaps from hPSCs. **(A)** Representative immunohistology staining of lung tissues from healthy control (CON) and PH patients against CD31. Right-hand panels show magnified views of the dashed regions in the left-hand images. Scale bars: 50 μm (main images), 20 μm (insets) **(B)** 3D-reconstructed images of lung samples described in (A), stained for CD31. Magnified views of the boxed regions highlight dilated capillaries (arrows). Scale bars: 50 μm; magnified images, 20 μm. **(C)** Dot plots show the capillary diameter in alveolar regions across groups (n=3 in each group). **(D-E)** Representative images of immunofluorescence staining for α-SMA or CD31 (green), CD34 (red) and nuclear DAPI staining (blue) of PH patients and CON lung tissues, bar=20 um. **(F)** Quantification of CD34^+^ endothelial cells in PH patients and CON lung tissues. **(G)** The mRNA level of aCaps markers (*APLN, EDNRB, HPGD* and *CAR4*) in whole lung tissues (n=3/group). **(H)** Schematic representation of the CRISPR/Cas9-mediated gene editing strategy used to insert a linker-EGFP construct into the 3′ region of the FOXF1 gene locus, preserving the integrity of the FOXF1 coding sequence and enabling the generation of a novel FOXF1-EGFP hPSCs. **(I)** Representative image shows the cell morphology and expression of EGFP in hPSCs after differentiation. **(J)** Representative image of FOXF1 and APLNR costaining or CD144 with APLN in Caps differentiated from hPSCs. **(K)** Flow cytometry reveals the distribution of gCap-like cells (APLNR⁺ FOXF1^+^) and aCap-like cells (APLN⁺) at different time points (n=3/group). **(L)** The mRNA level of gCaps marker (*CD34*), and aCaps markers (*APLN, EDNRB* and *HPGD*) at different stages in hPSCs from PH patients with BMPR2 mutation (n=3/group). All results were presented as mean±SEM. *: p<0.05, **: p<0.01, ***: p<0.001 and ****: p<0.0001 versus control and other groups as determined using t-test, one-way ANOVA or two-way ANOVA.

The obtacle to study Caps is lack of isolation and primary culture method due to the scarce of the recently identified cell types. We established an in vitro differentiation model to derive alveolar capillary like cells from hPSCs and examine the marker expression in the presence of BMPR2 mutation, the most frequently mutated gene in genetic form of PH^21^. Firstly, we generated FOXF1 (a gCap marker)-reporter human pluripotent stem cells (FOXF1-hPSCs) with CRISPR/Cas9-mediated knock-in of FOXF1-GFP^22^. This reporter system enables lineage tracing of gCap-like cells upon FOXF1 expression (Fig. 1, H). FOXF1-hPSCs were induced toward mesoderm differentiation, then the starting medium was replaced with capillary endothelial cell induction medium to Day 11. During differentiation, the CD34 levels of the cells progressively increased, and the expression of FOXF1 and APLNR was predominant on day 7 (D7), which was consistent with a gCap-like phenotype. By day 11 (D11), we observed a merger of APLN⁺ aCap-like cells that was accompanied by upregulation of *APLN*, *HPGD*, *TBX2*, and *EDNRB*, which indicated differentiated aCaps phenotypes (Fig. 1, I-K; and Fig. S3, A-D). Compared with control, HPAH-hPSCs with BMPR2 mutation showed increased expression of *CD34* (gCaps) at D7 and downregulated *APLN*, *EDNRB* and *HPGD* (aCaps) markers at D11 (Fig. 1, L). This result suggested reduced differentiation of aCaps in BMPR2 mutant hPSCs, it contributed to pulmonary capillary remodelling and dilation in PH patients

### Setting up a dynamic Capillary-Alveoli Micro-physiological System (CAMS) to recapitulate the two phase alteration under hypoxia

To help forming capillary network-like structure, we applied microfluidic chip with dynamic culture to test capillary integrity under PH, such as hypoxic condition. We established a Capillary-Alveoli Microphysiological System (CAMS) -intergrated vascularized alveoli organoid and planted into the microfluidic chip (Fig. 2, A). First, lung epithelial and stromal organoids were differentiated from hPSCs. Immunostaining revealed robust CDH1 expression in epithelial cells, accompanied by upregulated mRNA levels of *NKX2.1* and *SFTPC*, indicating successful epithelial differentiation. Stromal identity was confirmed by positive COL1 immunostaining and increased *COL3A1* expression (Fig. 2, B and C). Immunostaining for CD144 demonstrated that human pulmonary microvascular endothelial cell (hPMECs) formed a multilayered, interconnected microvascular network with lumen-like three-dimensional structures. Notably, a capillary network-like architecture was clearly observed in the dynamically perfused CAMS, with ECs closely associated with epithelial (CDH1⁺) and stromal (COL1⁺) organoids (Fig. 2, D and E).

**Figure 2.**
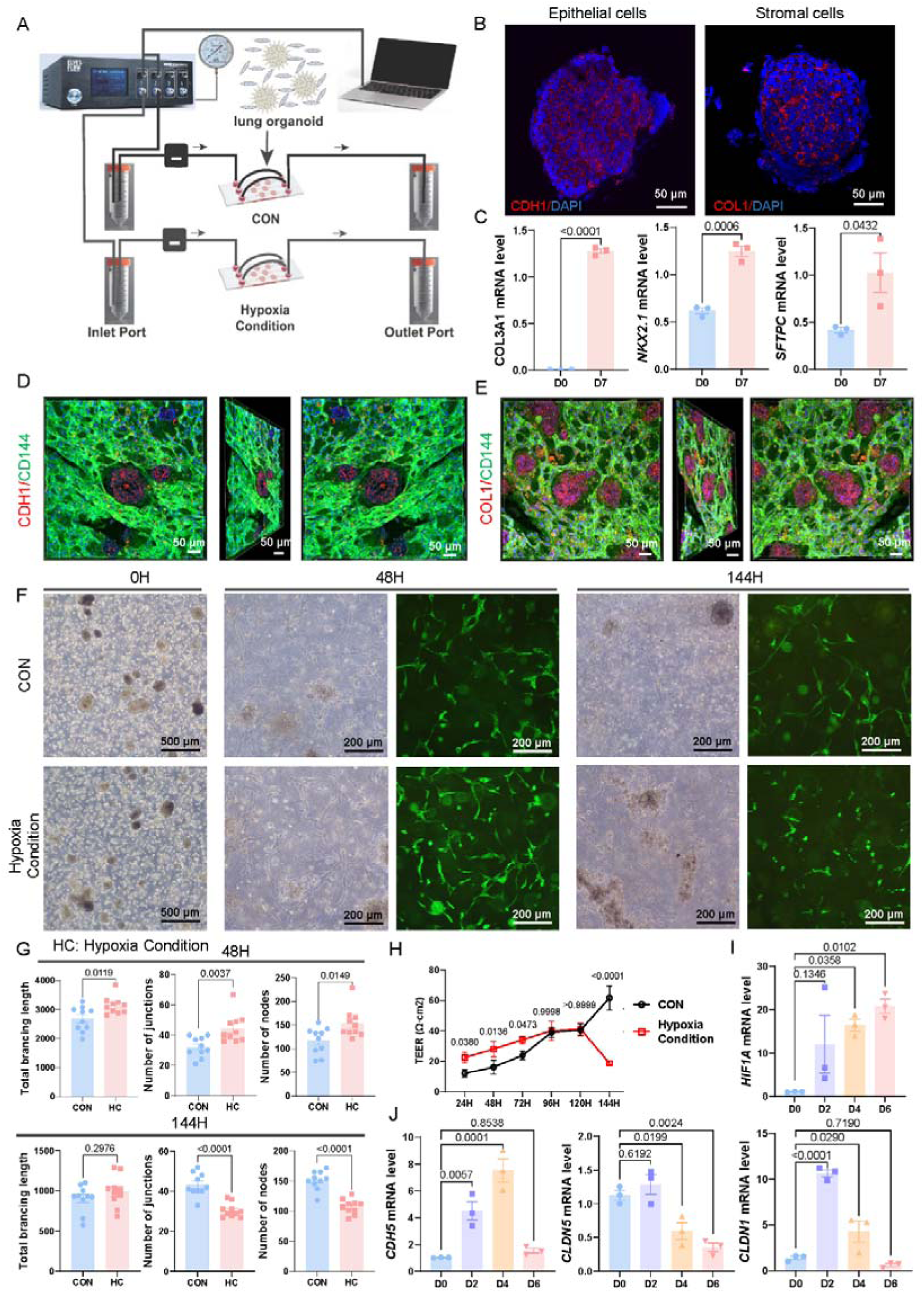
Setting up a dynamic Capillary-Alveoli Micro-physiological System (CAMS) to recapitulate the two phase alteration under hypoxia. **(A)** Schematic of constructing CAMS model. **(B)** Representative image of CDH1 or COL1 staining in cells differentiated from hPSCs. **(C)** The mRNA level of *COL3A1*, *NKX2.1* and *SFTPC* in cells mentioned in G. **(D-E)** 3D remodeling of lung organod. ECs (green) and CDH1 (red, D) or COL1 (red, E). **(F)** The bright-field ECs with autofluorescence of organoids at different time points. **(G)** Quantification the indexes of Total brancing length, Number of junctions and Number of nodes under hypoxia condition in different time points. **(H)** The electrical resistance, measured using the EVOM2 voltohmmeter, reflects the function of respiratory membrane. **(I-J)** The expression of *HIF1A*, *CDH5*, *CLDN5* and *CLDN1* in organoid model under hypoxia condition. All results are presented as mean±SEM. *: p<0.05, **: p<0.01, ***: p<0.001 and ****: p<0.0001 versus control and other groups as determined using t-test, one-way ANOVA or two-way ANOVA.

To model PH-associated hypoxia condition, CAMS was treated with CoCl₂ to induce chemical hypoxia^23–25^. Immunostaining and quantitative analysis of EGFP-transfected hPMECs at multiple time points revealed a response to hypoxia: short-term exposure enhanced endothelial tube formation, whereas prolonged hypoxia led to marked impairment of network integrity (Fig. 2, F). Quantification of total branching length, number of junctions, and number of nodes of Caps showed the significant increased tube maturation; whereas reduced endothelial tube formation under long term hypoxia exposure (Fig. 2, G; and Fig. S3, E). To further assess whether hypoxia compromised barrier integrity, trans-bilayer electrical resistance (TEER) was measured. Prolonged hypoxic treatment resulted in a significant decrease in TEER, indicating disruption of the endothelial-epithelial barrier structure (Fig. 2, H). At the molecular level, sustained upregulation of *HIF1A* confirmed successful induction of hypoxia, while a significant reduction in the expression of cell-cell junction molecular markers *CDH5, CLDN5, and CLDN1* suggested impaired barrier integrity during prolonged hypoxic exposure (Fig. 2, I and J). It implied that the alveolar-capillary network underwent a two-phase alteration under hypoxic conditions: a compensation stage followed by an uncompensation stage.

### Identification of elevated CD93 expression in CD34^+^APLNR^+^ gCaps of PH lungs

To understand how CD34^+^ ECs contribute to PH, we analysed previously published scRNA-seq data from lung samples from 3 PH patients and 6 healthy donors with even cell number^26^. After quality control, 33,540 single-cell transcriptomes were retained. On the basis of the expression of canonical marker genes, the cells were assigned to endothelial, epithelial, mesenchymal and immune compartments and further subdivided into subclusters (Fig. S4, A). In endothelial subclusters, pseudotime analysis showed that gCaps gave rise to aCaps as reported in lung development (Fig. 3, A and B; and Fig. S4, B) ^10^. This relationship was supported by lineage tracing in *Cd34-CreERT2; R26-tdTomato* mice, in which tdTomato^+^ (Cd34^+^) Aplnr^+^/Apln^+^ cells were detected, which indicated that Cd34^+^ gCaps generated aCaps subsets (Fig. S4, C). Consistent with impaired gCaps-to-aCaps transition, Ro/e analysis revealed that aCaps were depleted and gCaps were enriched in lung samples from PH patients compared with those from healthy controls (Fig. 3, C). These findings were corroborated by the quantification of APLNR^+^, APLN^+^, gCaps and aCaps populations in lung sections costained with CD34 and APLNR for gCaps and with CD31 and APLN for aCaps (Fig. S5).

**Figure 3.**
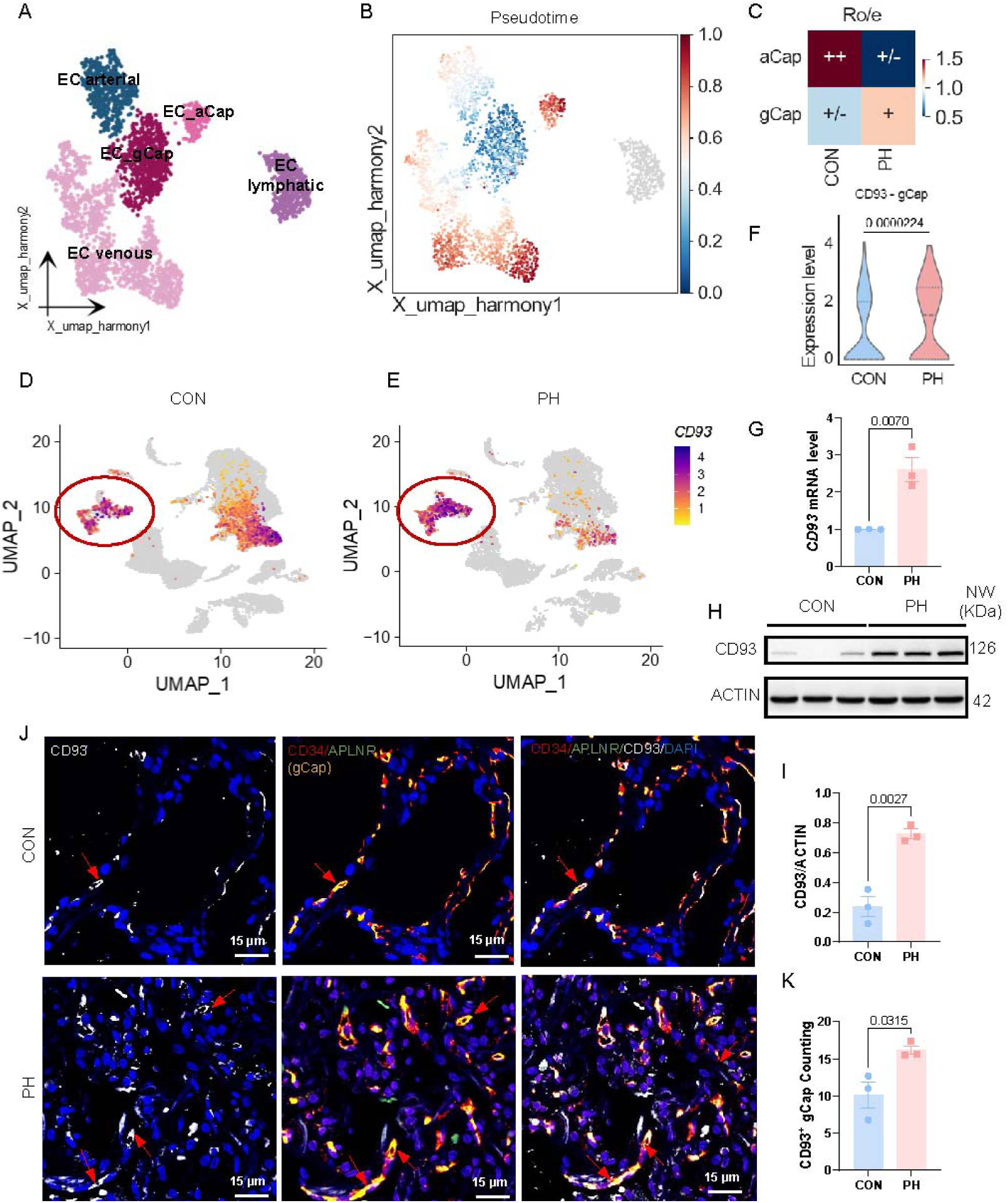
Identification of elevated CD93 expression in CD34^+^APLNR^+^ gCaps of PH lungs. **(A)** UMAP plot shows endothelial subpopulations from the integrated CON and PH single-cell RNA-seq dataset, colored by endothelial subtypes. **(B)** UMAP plot of endothelial cells colored by normalized diffusion pseudotime values (0-1 range). **(C)** Ro/e analysis reveals the relative enrichment or depletion of the two capillary endothelial subsets (gCaps and aCaps) in CON versus PH lungs. **(D**-**E)** UMAP feature plot illustrates CD93 expression in endothelial cells from CON and PH lungs. **(F)** Violin plot comparing CD93 expression in gCaps between CON and PH lungs. **(G)** The mRNA level of CD93 in lung lystate of PH patients and healthy CONs (n=3/group). **(H-I)** Representative Western blots and corresponding densitometric quantification demonstrate the expression of CD93 in patients’ lungs (n=3/group). **(J)** Representative images of immunofluorescence staining for CD93 (white) with gCaps markers: CD34 (red) and APLNR (green) in lung sections at different groups, bar=15 μm. **(K)** Quantification of CD93^+^ cells in gCaps. All results are presented as mean±SEM. *: p<0.05, **: p<0.01, ***: p<0.001 and ****: p<0.0001 versus control and other groups as determined using t-test.

To investigate the molecular basis of these changes, we performed DEG analysis of in gCaps from lung samples from control and PH patients and screened out significantly upregulated *CD93* which is involved in pathological angiogenesis ^27–31^. Analysis across all cell populations revealed that changes in *CD93* expression was most prominent in ECs; marked *CD93* upregulation occurred in gCaps and venous ECs, but minimal changes occurred in aCaps or arterial and lymphatic ECs (Fig. 3, D-F; and Fig. S6, A). Quantitative RT-PCR confirmed higher *CD93* mRNA levels in lung samples of PH patients than that of control donors (Fig. 3, G). At the protein level, western blot analysis confirmed CD93 upregulation in PH (Fig. 3, H and I). Immunofluorescence costaining for CD93 with CD34 and APLNR, with CD31, or with CD34 alone revealed increased numbers of CD93^+^ cells in the alveolar regions of lungs from PH patients (Fig. 3, J; and Fig. S6, B and C). Image quantification revealed increased densities of CD93^+^ cells among gCaps in lung samples from PH patients (Fig. 3, K). Overall, the marked increase in CD93 expression in the gCaps of patients with PH implied that CD93 Caps suppressed gCaps differentiation toward aCaps and contributed to PH.

### Upregulated Cd93 at remodeled capillary bed in Sugen/hypoxia rat lungs

Given the potential confounding effects of comorbidities in patient-derived samples, we next validated the differential expression of Cd93 using a well-established SuHx rat model under controlled experimental conditions (Fig. 4, A)^32^. The VEGFR2 inhibitor successfully induced key haemodynamic indices; specifically, SuHx rats exhibited significant increases in right ventricular systolic pressure (RVSP) and right ventricular hypertrophy, as assessed by the right ventricular hypertrophy index (RV/(LV+S)) (Fig. 4, B and C; and Fig. S7, A). Compared with control rats, SuHx rats also had a higher total pulmonary vascular resistance index (tPVRI) and lower cardiac output, cardiac index values, tricuspid annular plane systolic excursion (TAPSE), these indexes demonstrated that the rats developed PH.

**Figure 4.**
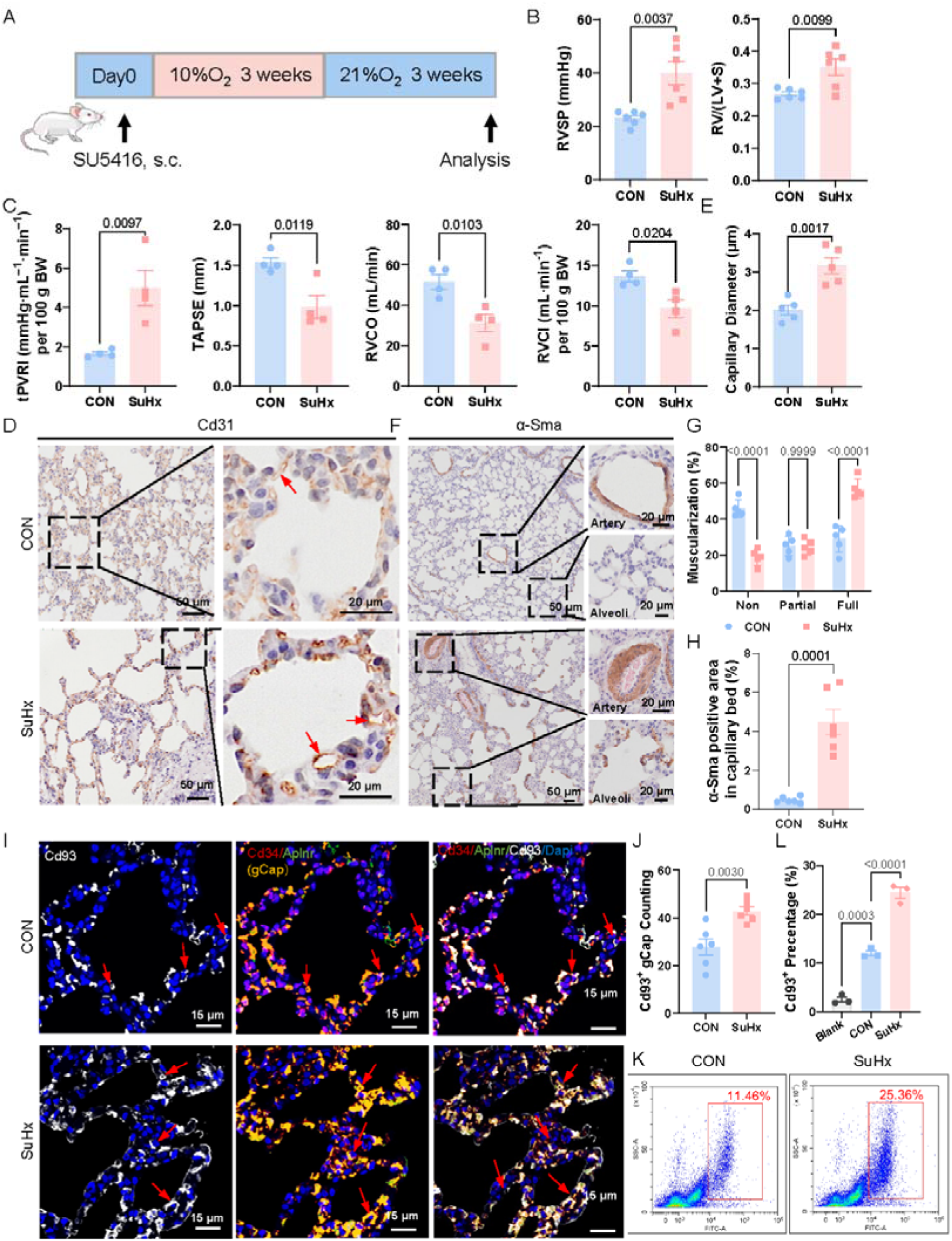
Upregulated Cd93 at remodeled capillary bed in Sugen/hypoxia rat lungs. **(A)** Schematic showing Sugen-hypoxia (SuHx) induced pulmonary hypertension in Sprague-Dawley (SD) rats (n=6/group). **(B-C)** Hemodynamic and cardiac function assessments. (B) Right ventricular systolic pressure (RVSP) and Fulton index [RV/(LV+S)] as an indicator of right ventricular hypertrophy (n=6/group). (C) Total pulmonary vascular resistance index (tPVRI = RVSP/RVCI), tricuspid annular plane systolic excursion (TAPSE), right ventricular cardiac output (RVCO) and right ventricular cardiac index (RVCI) (n=4/group) in different group. **(D-H)** Representative histological analysis of lung sections from control (CON) and SuHx rats. Cd31 staining reveals capillary dilation (D, quantified in E), while α-Sma staining highlights vascular remodeling and muscularization in alveolar capillaries (F-H) (n=6/group). **(I)** Representative immunofluorescence images show Cd93 expression co-localized with gCaps (Cd34⁺APLNR⁺) in SuHx lungs. Arrows indicate Cd93⁺Cd34⁺APLNR⁺ triple-positive cells. Magnified views correspond to boxed regions in the main images. Scale bars: 30 μm (main), 15 μm (insets); (n=6/group). **(J)** Quantification of CD34^+^ cells among gCaps populations in CON and SuHx lungs (n=6/group). **(K and L)** Flow cytometry analysis reveals the percentage of Cd93⁺ cells in lung tissue from CON and SuHx rats (n=3/group). All results are presented as mean±SEM. *: p<0.05, **: p<0.01, and ****: p<0.0001 versus control and other groups as determined using t-test or two-way ANOVA.

Histological analysis confirmed the pathological remodeling of the pulmonary vasculature. H&E staining (Fig. S7, B), Masson’s trichrome staining (Fig. S7, C and D) and elastin–van Gieson (EVG) staining (Fig. S7, E) exhibited vascular remodeling and collagen deposition in the lungs of SuHx rats. Capillary bed dilation was detected by IHC staining for Cd31, which confirmed that the capillary lumen diameter was greater in alveolar regions (Fig. 4, D and E; and Fig. S7, F). Furthermore, α-Sma staining revealed enhanced muscularization of small arteries and alveolar capillaries in the lungs of SuHx rats, with quantitative analysis confirming a significant increase in wall thickness (Fig. 4, F-H; and Fig. S7, G).

Next, IHC confirmed that Cd93 expression was elevated across pulmonary vessels (Fig. S7, H). We showed that Cd93 expression increased in Cd34^+^Aplnr^+^ double-positive gCaps (Fig. 4, I) and in Cd31^+^ (Fig. S7, I) or Cd34^+^ cells (Fig. S7, J). Quantitative analysis revealed a significant increase in the density of Cd93⁺ cells among gCaps in SuHx rats compared with controls (Fig. 4, J). Flow cytometry analysis corroborated these findings by revealing a significant increase in the proportion of Cd93⁺ cells in SuHx rats (Fig. 4, K and L). Importantly, similar upregulation of Cd93⁺ cells among total lung cells and Cd34^+^ cells was observed in an independent SuHx model using *Cd34*-lineage tracing mice (Fig. S8, A-C), which supports a conserved role of Cd93 in PH pathogenesis. Taken together, these data confirmed the upregulation of Cd93 expression among gCaps in the experimental PH model, validating its association with pulmonary capillary remodeling.

### Deletion of Cd93 in Cd34⁺ cells attenuate pulmonary capillary remodeling and PH by rescued differentiation toward aCaps and vice versa

To investigate the functional role of CD93 in the progression of PH, we generated a conditional *Cd93* knockout mouse model. we employed a *Cd34*-CreER^T2^; R26-tdTomato; *Cd93^flox/flox^ (Cd93*^Δ*Cd*34^) mouse model, in which *Cd93* was selectively deleted in Cd34⁺ gCaps. Under SuHx conditions, *Cd93*^Δ*Cd*34^ mice exhibited a significant reduction in RVSP but no significant reduction in RV/(LV+S) compared with their *Cd93^f/f^* littermates (Fig. 5, A; and Fig. S8, B).

**Figure 5.**
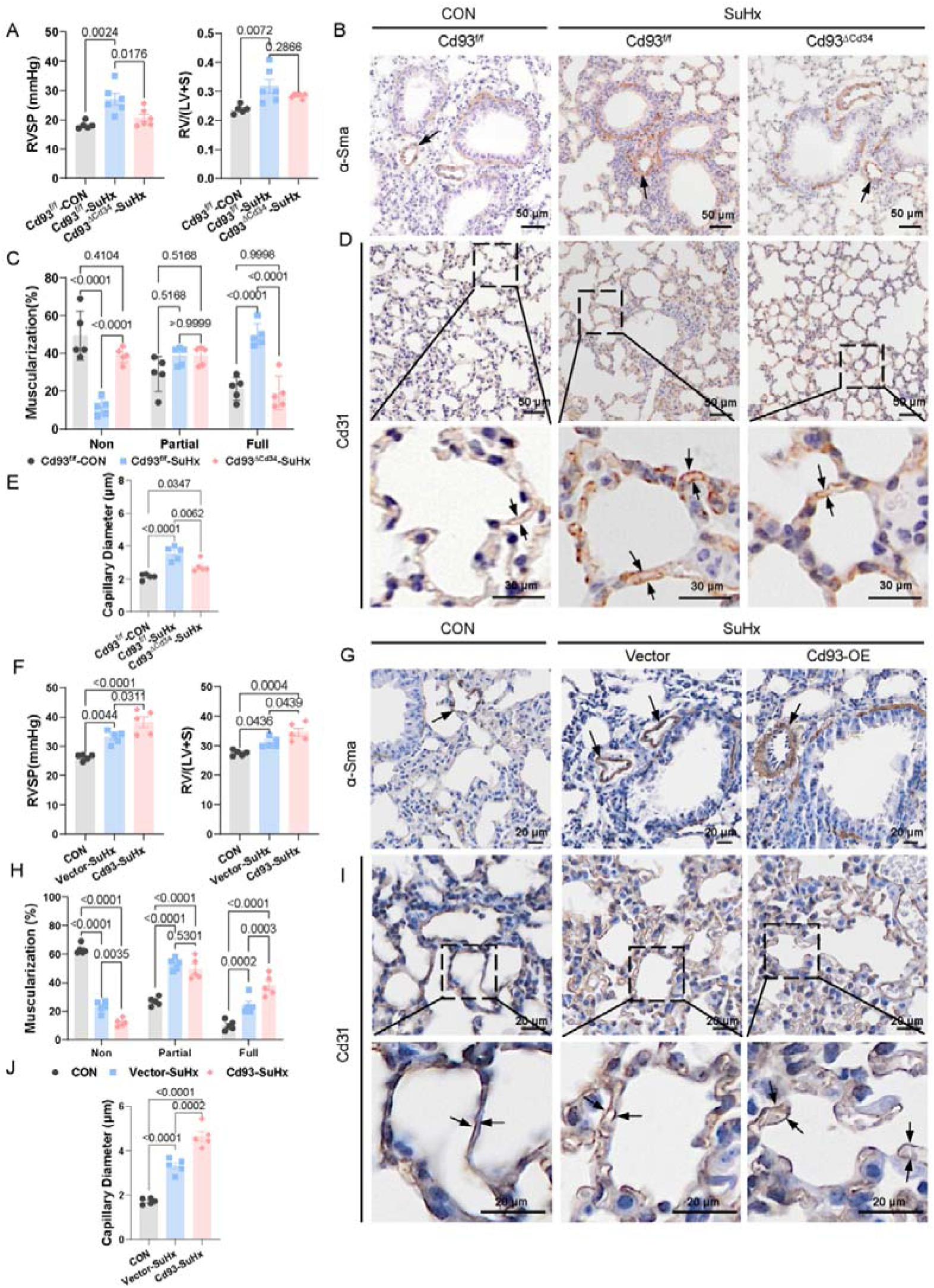
Deletion of Cd93 in Cd34⁺ cells attenuate pulmonary capillary remodeling and PH by rescued differentiation toward aCaps and vice versa. **(A)** Hemodynamic measurements show RVSP and RV/(LV+S) in Cd93^ΔCd34^-SuHx mouse lungs compared to Cd93^f/f^-SuHx and controls (n=5/group). **(B-C)** α-Sma staining and quantification illustrate vascular remodeling in each group. Scale bars: 50 μm, (n=5/group). **(D-E)** Cd31-IHC staining and quantification show changes in capillary dilation between groups, Scale bars: 50 μm (main), 30 μm (insets), (n=30/group). **(F)** Hemodynamic measurements reveal RVSP and RV/(LV+S) in Cd93-SuHx mouse lungs compared to Vector-SuHx and controls (n=5/group). **(G-H)** α-Sma staining and it’s quantification demonstrate vascular remodeling in each group. Scale bars: 20 μm, (n=5/group). **(I-J)** Cd31-IHC staining and quantification illustrated changes in capillary dilation between groups, Scale bars: 20 μm, (n=5/group). All results are presented as mean±SEM. *: p<0.05, **: p<0.01, and ****: p<0.0001 versus control and other groups as determined using one-way ANOVA or two-way ANOVA.

H&E staining and subsequent quantification revealed reduced vascular wall thickening in *Cd93*^Δ*Cd*34^ mice compared with controls (Fig. S8, D and E). Consistent with these observations, α-Sma immunohistochemistry revealed decreased muscularization of pulmonary vessels (Fig. 5, B and C). Additionally, Cd31 staining revealed improved capillary architecture with decreased capillary dilation in *Cd93*^Δ*Cd*34^ mice under SuHx conditions (Fig. 5, D and E), which suggests partial restoration of vascular homeostasis. Then, we ovexpressed Cd93 in lung tissues through intratracheal instillation of lentivirus and detected enhanced RVSP and RVHI indexes (Fig. 5, F). α-Sma and CD31 staining showed increased Capillary remodelling vice versa (Fig. 5, G-J).

To test whether *Cd93* affects capillary ECs differentiation, we performed dual immunofluorescence using tdTomato (Cd34) and Apln (aCaps marker). These analyses revealed that the conditional deletion of *Cd93* in Cd34⁺ cells reversed the SuHx-induced reduction in Apln⁺ cell populations (Fig. S8, F), indicating a rescue of gCaps-to-aCaps differentiation. Targeted deletion of *Cd93* in Cd34⁺ cells rescued Apln^+^ cells, partially restored capillary identity and alleviated pulmonary vascular remodeling, which suggests that Cd93 is a brake for gCaps differentiate into aCaps in PH. Taken together, these findings demonstrated that *Cd93* played an important role in the formation of aCaps.

### CD93 suppresses APLN expression through activation of SMAD2/3 and reduced capillary endothelium integrity

Our findings above strongly suggest that gCaps differentiation is regulated by CD93, however, the signalling pathway downstream of CD93 is poorly understood. Since hPMECs are ECs sorted from the human alveolar region. We first investigated this signalling in a CD93-overexpressing hPMEC line generated using a lentiviral delivery system (pLVX-CD93) and in a CD93-targeting siRNA treated hPMEC line with CD93 expression knockdown (Fig. 6, A-D).

**Figure 6.**
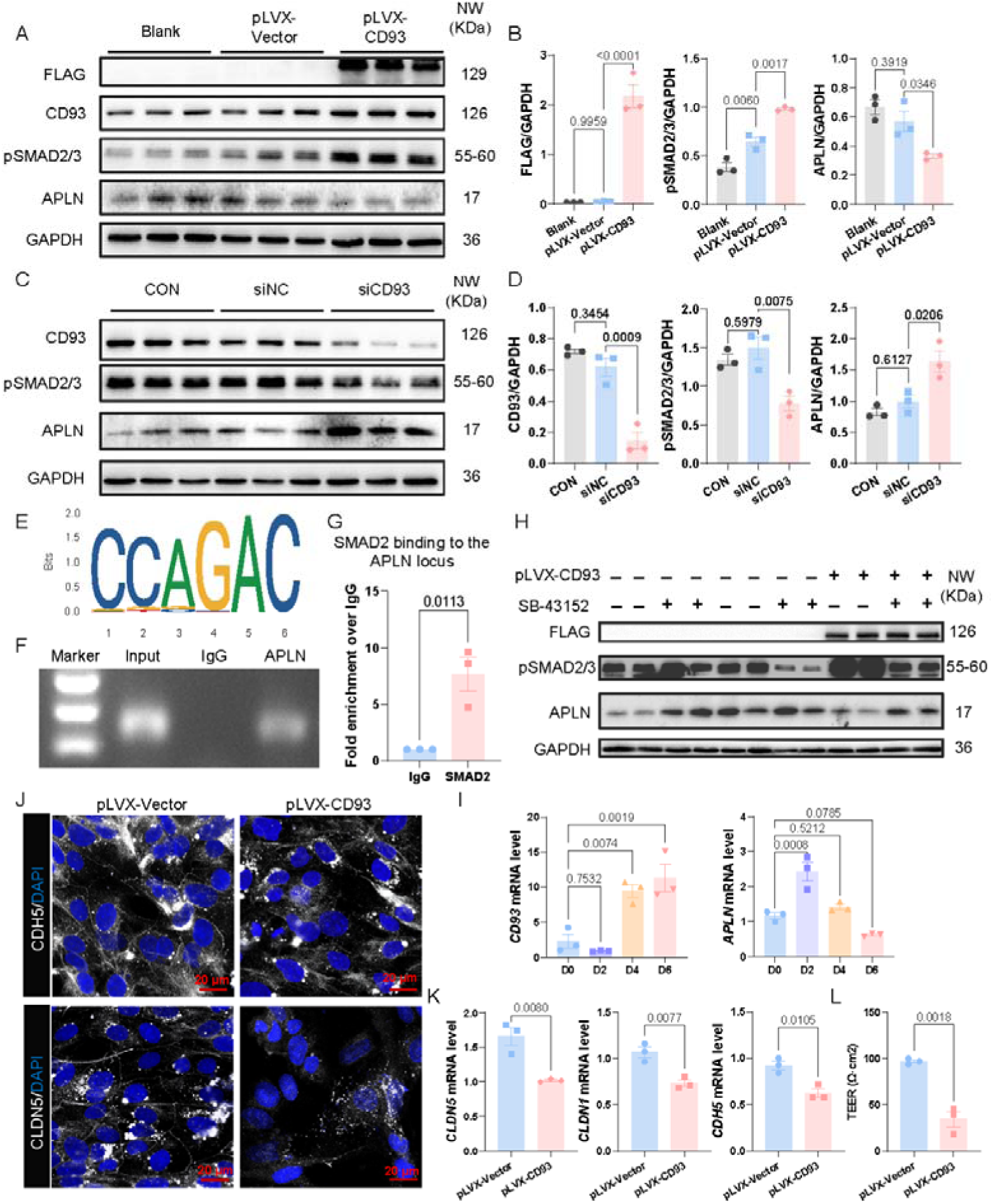
CD93 suppresses APLN expression through activation of SMAD2/3 and reduced capillary endothelium integrity. (A-B) Representative Western blots and corresponding densitometric quantification (n=3/group) show the expression of CD93 and pSMAD2/3 and APLN in pLVX-CD93 group. **(C-D)** Immunoblotting and densitometric quantification in hPMECs transfected with siCD93. **(E)** Predicted DNA binding site motif of SMAD2 in the APLN promoter region. **(F-G)** The APLN promoter region in hPMECs pulled down with an SMAD2 antibody was detected by chromatin immunoprecipitation (ChIP)-PCR (F) and ChIP-qPCR (G) n=3 in panel. **(H)** Immunoblotting of FLAG, P-SMAD2/3 and APLN in hPMECs, transfected with pLVX-CD93 vector, treatment with SB-431542 cells probed with the indicated antibodies. **(I)** The expression of *CD93* and *APLN* in organoid model under hypoxia condition in different time points (n=3/group). **(J)** Representative images of immunofluorescence staining for CDH5 and CLDN5 (white) in PLVX-Vector and PLVX-CD93 organoid models from CAMS, bar=20 μm. **(K)** The mRNA level of junction markers: CLDN5, CLDN1 and CDH5 in CAMS. **(L)** The TEER measured using the EVOM2 voltohmmeter in PLVX-Vector and PLVX-CD93 organoid models (n=3/group). All results are presented as mean±SEM. *: p<0.05, **: p<0.01, ***: p<0.001 and ****: p<0.0001 versus control and other groups as determined using one-way ANOVA.

Given that the signaling pathway downstream of CD93 being unknown and that CD93 knockout mice exhibited reduced levels of phosphorylated SMAD2/3^32, 33^, we hypothesized that CD93 might act through SMAD2/3 signalling in PH. Our findings demonstrated that CD93 overexpression significantly increased the levels of phosphorylated SMAD2/3 (pSMAD2/3) and markedly decreased the levels of the aCaps marker APLN at both the mRNA and protein levels (Fig. 6, A and B; and Fig. S9, A). Conversely, CD93 knockdown reduced SMAD2/3 activation but concomitantly increased APLN expression in hPMECs at both the both the gene and protein levels (Fig. 6, C and D; and Fig. S9, B).

To examine whether SMAD2/3 can regulate the expression of APLN directly, we predicted that the APLN promoter region contains the SMAD2 binding site sequence (AGAC) by searching JASPAR (http://jaspar.genereg.net) (Fig. 6, E). A chromatin immunoprecipitation-PCR assay revealed that the binding of SMAD2 to the APLN promoter region (Fig. 6, F and G). SB-431542, an inhibitor ALK4/5/7 reduced pSMAD2/3 activity increased APLN expression (Fig. S9, C and D), This result indicated that reduced pSMAD2/3 activation promoted APLN expression in hPMECs.

To assess whether CD93 regulated APLN primarily through SMAD2/3 activation, we treated CD93-overexpressing hPMECs with SB-431542(Fig. 6, H). Notably, the inhibition of SMAD2/3 restored APLN expression in CD93-overexpressing cells, which indicated that the suppressive effect of CD93 on APLN was SMAD2/3 dependent. We also confirmed in CAMS hypoxia increased CD93 expression with reduced APLN concomitantly (Fig. 6, I).

To demonstrate the interference of high level of CD93 at the endothelium junction formation, we transducted pLVX-CD93 into H9 human PSCs and differentiated into aCaps, the staining with CDH5 and CLDN5 showed reduced expression of both protein at the junction of Capillary ECs, which consistent with the mRNA level of CLDN5, CLDN1 and CDH5 (Fig. 6, J and K). The TEER result displayed that CD93 damages the capillary endothelial integrity (Fig. 6, L). Collectively, our data revealed that CD93 acts through the SMAD2/3 activation to repress APLN expression and contribute to the loss of integrity of capillary endothelium.

### APLNR agonists enhance the network formation of Capillary cells to prevent and reverse PH in rats without affecting heart hypertrophy

To find the potential target in CD93-pSMAD2/3-APLN signalling, we analysed the expression patterns of *CD93* and *APLN* in scRNA-seq datasets from human lung tissues (https://www.immunesinglecell.org). Our analysis revealed that *CD93* was broadly expressed across multiple ECs and immune cell subtypes, whereas *APLN* expression was selectively enriched in aCaps (Fig. S10, A and B). So we selectively targeted CD93’s downstream effector APLN to improve its binding with APLNR expressing gCaps and promote differentiation into functional aCaps in PH. Therefore, we evaluated the therapeutic potential of APLN with in vitro and in vivo PH models. However, traditional APLN peptides cannot be used therapeutically because they induce cardiac hypertrophy due to their β-arrestin-biased signalling properties^34^. Therefore, we instead tested two functionally optimized APLNR agonists/APLN analgues, WN353 and WN561, that were engineered in previous work to provide therapeutic efficacy while minimizing adverse effects^34^. To assess the efficacy of these agonists in rescuing PH-associated phenotypes, we first assessed them in complementary in vitro *BMPR2* deficient ECs for modelling PH in vitro: (1) hPSCs knockdown with *BMPR2* and differentiate into ECs. (2) hPMECs transfected with siBMPR2. Quantitative mRNA expression analysis demonstrated that both WN353 and WN561 significantly attenuated the BMPR2 deficiency upregulated expression of proinflammatory cytokines (*TNF-*α*, CCL5,* and *IL-6*) and endothelial-to-mesenchymal transition (EndoMT) markers (α*-SMA* and *CDH5*) (Fig. S10, C-G). The improvements in these indicators suggested that supplement of APLN inhibited key pathogenic pathways of PH in vitro.

Next, we evaluated the in vivo therapeutic efficacy of APLN and its analogues (WN353, and WN561) using a preventive protocol in a SuHx rat model (Fig. S11, A). Among these peptides, WN353 exhibited the most robust protective effects and produced the most significant reductions in RVSP and RV/(LV+S) (Fig. S11, B). Histopathological assessment further revealed that both WN353 and APLN markedly attenuated vascular remodeling across different vessel diameters, whereas WN561 had minimal effects (Fig. S11, C and D).

To assess the therapeutic potential of APLNR agonists in established PH, we conducted intervention studies in SuHx rats and compared the effects of WN353 and native APLN with those of sildenafil, a phosphodiesterase-5 inhibitor (PDE5i) that is commonly used to treat PH (Fig. 7, A). WN353 treatment significantly reduced RVSP and tPVRI , whereas both sildenafil and APLN had only modest or negligible therapeutic effects (Fig. 7, B). Notably, treatment with sildenafil, APLN or WN353 did not significantly improve the RV/(LV+S) which was used to evaluate the right heart function; however, it was markedly lower in the WN353 group than in the APLN group (Fig. S12, A). Histopathological assessments using H&E and α-Sma staining revealed that WN353 and sildenafil, but not native APLN, significantly reversed pulmonary vascular remodeling and vessel wall thickening (Fig. 7, C-F; and Fig. S12, B-D). Overall, these findings highlighted WN353 as a promising therapeutic agent capable of preventing and reversing pathological changes in experimental PH models

**Figure 7.**
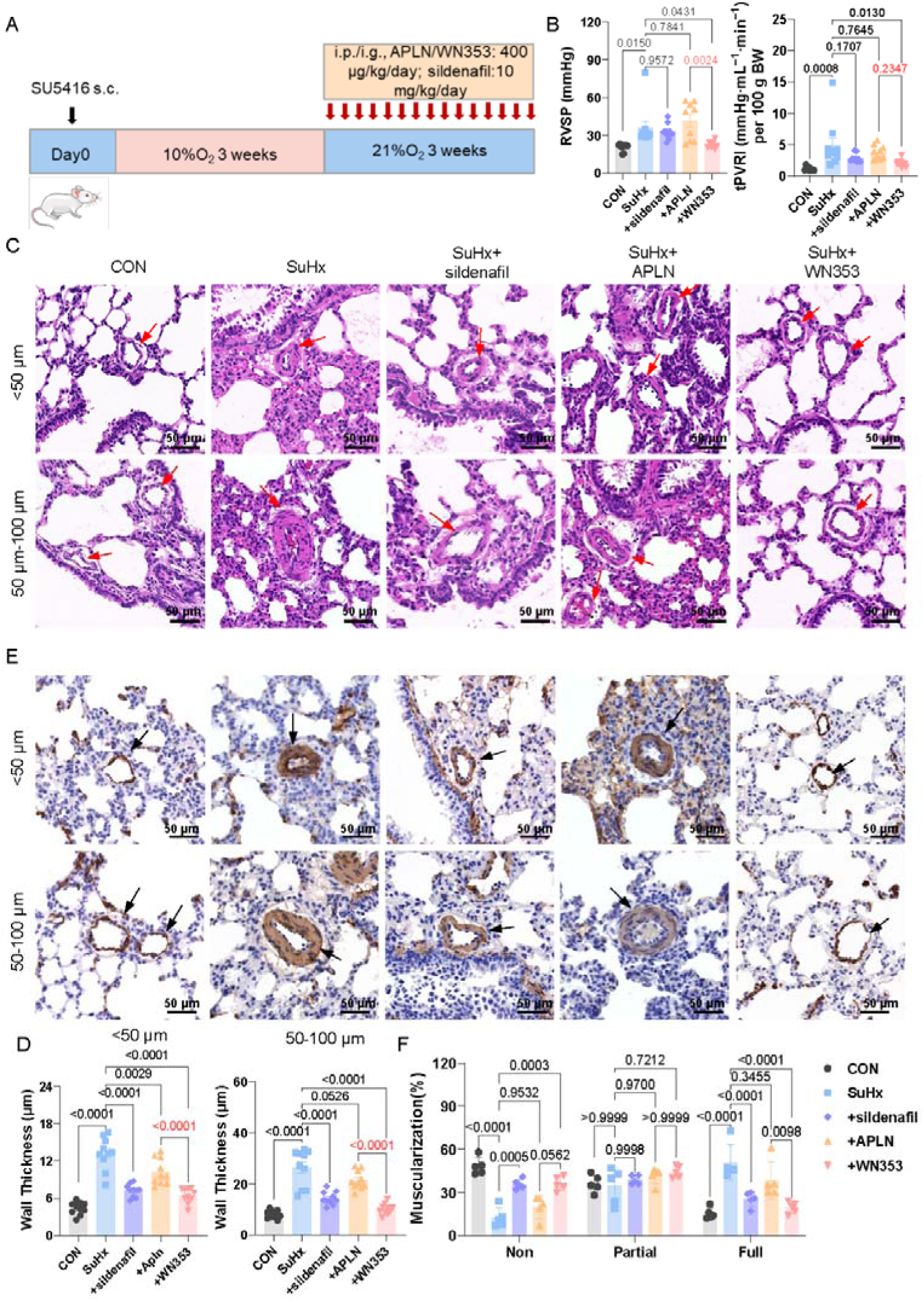
APLNR agonists enhance the network formation of Capillary cells to prevent and reverse PH in rats without affecting heart hypertrophy. **(A)** Schematic illustration of the experimental design and pharmacological treatment regimen in the SuHx model of PH using SD rats (n=10/groups). **(B)** Hemodynamic assessment of RVSP, tPVRI, across treatment groups. **(C-D)** Representative H&E staining and corresponding quantification of pulmonary vessels with varying luminal diameters (<50 μm and 50 μm-100μm), highlighting differences in vascular wall architecture among treatment groups. **(E-F)** α-Sma staining and quantification demonstrate the extent of vascular remodeling and medial wall thickness. Sildenafil and WN353 treatments significantly reduce vascular remodeling (n=10/group), bar=50 μm. All results are presented as mean±SEM. *: p<0.05, **: p<0.01, ***: p<0.001 and ****: P<0.0001 versus control and other groups as determined using one-way ANOVA or two-way ANOVA.

### APLNR agonists promote the regeneration of aCaps and alleviate pulmonary capillary bed remodelling

To determine whether APLNR agonists promoted Caps regeneration and alleviated pulmonary capillary remodeling, we assessed capillary diameter in alveolar regions following treatment (Fig.8, A). Sildenafil, APLN and WN353 each significantly reduced capillary dilation, and WN353 had the strongest effect (Fig. 8, B). Additionally, qRT-PCR analysis of lung tissue revealed that WN353, but not sildenafil or native APLN, significantly upregulated the expression of aCaps associated markers, including *Ednrb, Car4* and *Tbx2,* at the mRNA level (Fig. 8, C). These results suggested that WN353 attenuated capillary dilation in vivo and promoted the restoration of aCaps differentiation.

**Figure 8.**
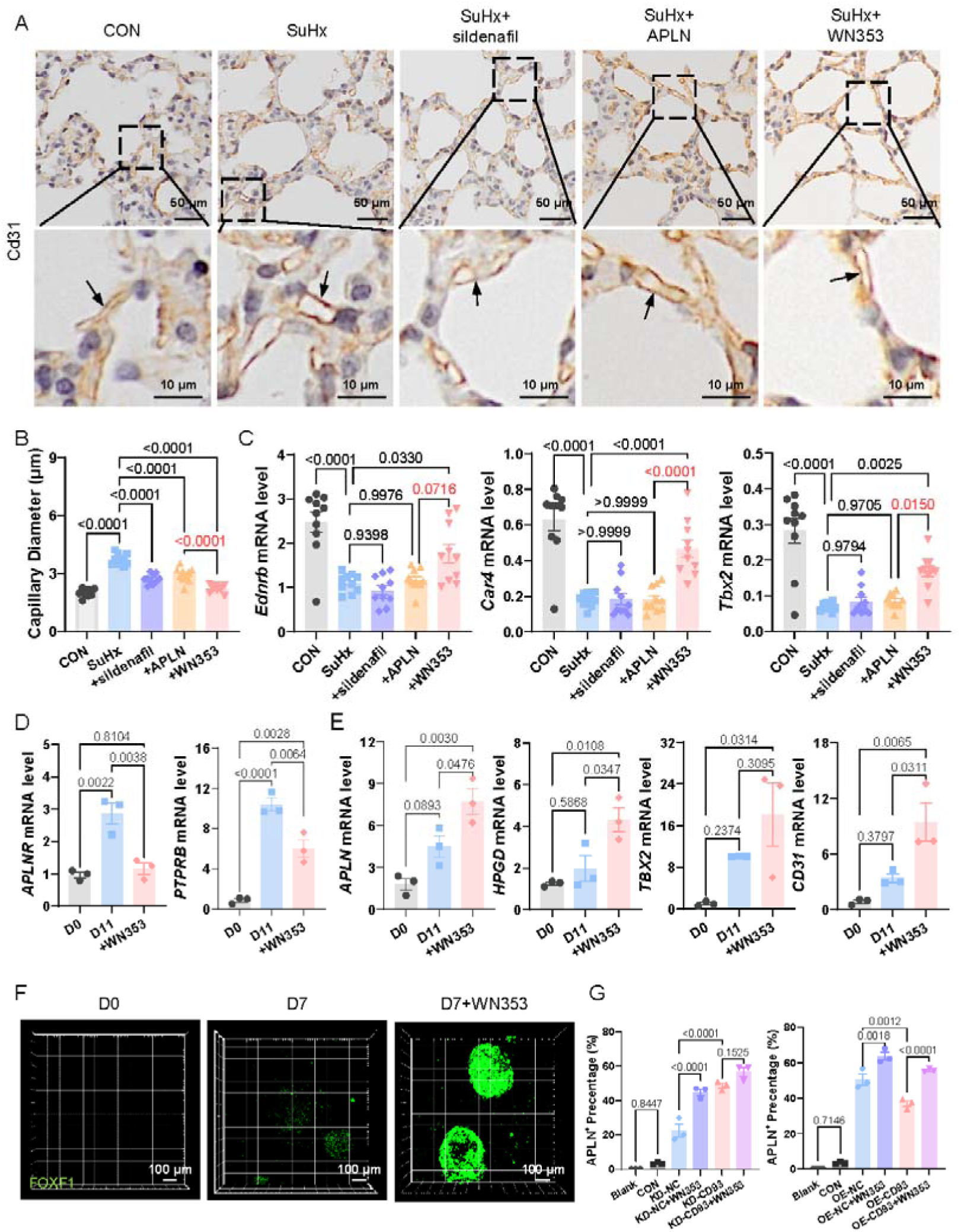
APLNR agonists promote the regeneration of aCaps and alleviate pulmonary capillary bed remodeling. (A-B) Cd31 staining and quantification demonstrate the diameter of capillary in alveoli region (n=10/group). Images on the below are magnified views of the dashed square in the up-hand images, bar=50 μm, bar=10 um. **(C)** The mRNA level of aCaps markers (*Ednrb, Car4* and *Tbx2*) in whole lung lystate at different groups. **(D-E)** qRT-PCR shows the expression of gCaps (*APLNR* and *PTPRB*) and aCaps (*APLN*, *HPGD* and *TBX2)* and *CD31* at D11 in supplement with WN353 during differentiation from gCap-like cells to aCap-like cells. **(F)** Representative image reveals the intensity of EGFP at D7 with the supplement of WN353. Scale bars: 100 μm. **(G)** Flow cytometry illustrates the percentage of aCap like cells, differentiated from hPSCs (pLVX-CD93 and CD93-KD cell lines with supplement of WN353, (n=3 /group). All results are presented as mean±SEM. *: p<0.05, **: p<0.01, ***: p<0.001 and ****: P<0.0001 versus control and other groups as determined using one-way ANOVA.

To further assess the role of WN353 in capillary cell differentiation in vitro, we incubated FOXF1-hPSCs with WN353-supplemented differentiation medium. WN353 treatment significantly reduced the expression of gCaps markers (*APLNR* and *PTPRB*) (Fig. 8, D). Concurrently, it increased the expression of aCaps markers (*APLN, HPGD* and *TBX2*) and the EC marker CD31 (Fig. 8, E). Consistent with these findings, autofluorescence imaging of FOXF1 on D7 revealed increased EGFP intensity and a higher percentage of EGFP⁺ cells following WN353 treatment (Fig. 8, F).

To test whether WN353 rescued gCaps-to-aCaps differentiation under PH conditions, we added WN353 at the gCaps-to-aCaps transition stage (from D7-D11) during pLVX-CD93 and CD93-KD FOXF1-hPSC differentiation. Flow cytometry analysis demonstrated that WN353 significantly increased the proportion of aCaps in both lines (Fig. 8, G), which indicated that WN353 could overcome CD93-mediated suppression while promoting aCaps differentiation. Taken together, these findings highlight WN353 as a potent modulator of capillary cell plasticity that is capable of restoring aCaps differentiation in vivo and driving gCaps-to-aCaps differentiation in vitro.

## Discussion

This study aimed to elucidate the role of Caps alteration and its underlying regulatory mechanism in PH. We employed an integrated approach involving a novel Capillary-Alveoli Micro-physiological System (CAMS) with in vitro hPSCs-aCaps differentiation strategy, high-resolution confocal microscopy, scRNA-seq on lineage tracing mice to identify the molecular cascade underlying capillary cell differentiation. We Knockdown Cd93 in Cd34^+^ gCaps of PH mice to examine the role of Cd93, together with a CHIP assay to confirm the transcription repression of APLN by pSMAD2/3, which is a non-canonical pSMAD2 activation and suggested to the regulatory role of the CD93-pSMAD2/3-APLN signaling axis in gCaps differentiation. Here, we report previously unrevealed alveolar capillary bed alteration in PH patients and animal models. It was characterized by pronounced capillary dilation and aberrant muscularization within the alveolar capillary bed. Importantly, the differentiation of gCaps into specialized aCaps was markedly suppressed by elevated expression of CD93. This disruption in capillary cell differentiation plays a critical role in the pathogenesis of PH by preventing the restoration of normal alveolar capillary function.

Although it was reported that FOXF1 loss-of-function mutation in gCaps resulted in defective angiogenesis and a reduced number of alveolar capillaries in Alveolar Capillary Dysplasia due to disrupted BMP9/ACVRL1 signaling ^22^, the possible role of Caps in PH was only mentioned at thickened arterial wall with proliferative ECs in neointima of artery. Liu B et al showed that gCaps can transdifferentiate into PAECs through HIF-2α/Notch4 signaling, leading to the muscularization of the pulmonary artery^16^. The dilated pulmonary capillary beds haven’t been formally documented in PH study since the limited methodology available until we set up CAMS incorporated with capillary endothelial cells, epithelial cells and stromal cells mixed organoid, which provided three major layers of respiratory membrane. We found short period(24H) hypoxic condition increased formation of capillary networks within the complex, whereas long term(120H) CoCl_2_ treatment reduced barrier integrity.

To determine the underlying molecular mechanism in dysfunctional capillary, we identified CD93 as a critical modulator in gCaps among the DEGs with reduced aCaps from scRNA-seq analysis on PH patient lung tissues. CD93 expression was significantly elevated in lung tissues from PH patients. Importantly, Cd93^-/-^ mice exhibited weaker Smad2/3 activation, supporting a functional role for Cd93 in maintaining the activity of this key signalling cascade^33^. APLN and SMAD2/3 expression also responded to TGF-β1 stimulation^35^ ^36^, thus implies the possible crosstalk of CD93 with TGF-β1 receptor at cell membrane. Taken together, these findings support a model in which CD93 acts upstream of the pSMAD2/3-APLN axis to drive gCaps-to-aCaps differentiation, and the disruption of this pathway contributes to defective capillary network formation in PH.

To investigate the regulation of gCaps-to-aCaps differentiation, we established an hPSCs-derived capillary cell differentiation protocol and successfully generated aCaps with mature marker expression. In vitro experiments demonstrated that APLN can increase gCaps-to-aCaps cell differentiation. However, native APLN peptides are known to have undesirable side effects, including cardiac hypertrophy, which limits their therapeutic use. In a previous study, we employed β-arrestin-biased APLNR agonists, including WN353 and WN561, which preferentially activate downstream signalling pathways without inducing hypertrophic responses. In both preventive and therapeutic models, administration of the APLNR agonist WN353 not only prevented but also reversed disease progression in a PH rat model. Importantly, WN353 treatment also enhanced differentiation from gCaps to aCaps. These findings demonstrated that augmenting aCaps differentiation via the APLNR agonist WN353 reversed pulmonary vascular remodeling. Overall, this novel strategy for PH treatment overcame a previous barrier to treatment posed by native APLN and preferentially acted on endothelial dysfunction.

One limitation of the current study lies in the assessment of the suppressed gCaps-to-aCaps differentiation process using scRNA-seq analysis. Ideally, single-cell transcriptomic profiling should be performed on lung tissues collected across a spectrum of PH disease severity. This would allow for the identification of trends in gCaps-to-aCaps differentiation during disease progression and help determine whether and when this process is inhibited. However, owing to clinical constraints, we were unable to obtain lung tissue specimens representing different disease stages. Another limitation concerned is to determine the origin of capillary dilation, one must consider its unique structure. Unlike muscular vessels, capillaries lack smooth muscle and cannot actively constrict. Therefore, observed dilation is often a passive response to increased intravascular pressure. Alternatively, it can result from pathological wall remodeling, such as that induced by inflammatory factors leaking from the plasma into the interstitium. Another possibility is the mutation contributes to the weakened support of stromal cells and thereby increased capillary diameter. Our CAMS system provides a platform for further investigating these issues.

In summary, our study identified the presence of alveolar-capillary remodeling and suppressed gCaps-to-aCaps differentiation in PH, and we set up CAMS model to study capillary alteration under pathophysiological condition. Using this system, together with animal models we identified the CD93 as a key regulatory gene that controlled capillary network formation, and we demonstrated that the G protein-biased APLN analogue WN353 effectively promoted gCaps-to-aCaps differentiation, with therapeutic effects on PH models in vitro and in vivo. Further research with CAMS model, we could examine the function of respiratory membrane in human genetic or inflammatory vascular diseases. WN353 was shown to prevent and reverse PH pathology, thus highlighting the therapeutic potential of modulating gCaps-to-aCaps differentiation as a novel strategy for pulmonary vascular disease.

## Materials and Methods

### Animals

All animal procedures were approved by the Institutional Animal Care and Use Committee (IACUC) of Zhejiang University School of Medicine and conducted in accordance with institutional guidelines for the ethical care and use of laboratory animals with Ethics code: ZJU20210036.

### Mouse generation and breeding

Mice with an Cd34-specific deletion of Cd93 (generated by breeding *Cd93*^fl/fl^ mice with *Cd34*-Cre mice. Cd34-CreER^T2^ and Cd93^fl/fl^ mice were provided by Dr. Chen Dong (Tsinghua University) and Dr. Qingbo Xu (Zhejiang University), respectively. Genomic DNA was extracted from mice tail tissue by using One Step Mouse Genotyping Kit (Vazyme, Cat. PD101-01).

### SuHx animal model

Male Sprague Dawley rats (160 to 180 g, GemPharmatech) or Cd93^ΔCd34^ mice (8-week-old) were administered to the SU5416/hypoxia (SuHx) model to induce PH. For rats received a single subcutaneous injection of SU5416 (20 mg/kg; Aladdin, Cat. 204005-46-9), followed by exposure to 10% O₂ hypoxia for 21 days and then returned to normoxia for 21 days. For lineage tracing mice, they were administered tamoxifen to induce recombination, followed by SU5416 once a week for 3 weeks and exposure to chronic hypoxia for 3 weeks, after which the mice were returned to normoxic conditions for an additional 3 weeks to induce PH.

For hemodynamic assessments, animals were anesthetized via intraperitoneal injection of sodium pentobarbital (1 or 2% in 0.9% saline). Right ventricular systolic pressure (RVSP) was acquired by a Millar catheter (Smith Medical). Right ventricular hypertrophy index was quantified by calculating the Fulton index. Following euthanasia, the right lung was fixed in 4% paraformaldehyde (PFA) for histological analysis.

For prevention experiment, at day0 of the SuHx model, rats were randomly assigned to five groups: One group was normal control group (injected with H_2_O), other was normal model group (injected with H_2_O), the remaining three groups were prevention groups (injected with APLN, WN353 or WN561, separately). The compounds APLN, WN353, and WN561 were gifts from Dr. Yan Zhang (Zhejiang University). Peptides were administered every other day for six weeks during disease development.

For reversal experiment, three out of the five groups were the treatment groups (injected with sildenafil, APLN or WN353, separately). After return to normoxia, these rats underwent a three-week course of daily drug administration.

### Cell lines

Human pulmonary microvascular endothelial cells (hPMECs) were obtained from (OLOGOBIO, China) and cultured in complete endothelial cell medium (ECM; ScienCell, Cat. 1001), supplemented with endothelial cell growth supplement (ECGS; Cat. #1052), 5% fetal bovine serum (FBS; Gibco, Cat. #0025), and penicillin/streptomycin (Pen/Strep; Cat. #0503). HEK293T cells were cultured following the standard protocols provided by the

American Type Culture Collection (ATCC). HPSCs were maintained in mTeSR1 medium (STEMCELL Technologies, Cat. 85850) under feeder-free conditions, according to the manufacturer’s instructions. Cells were maintained at 37 °C in a humidified incubator with 21% O₂ and 5% CO₂. The culture medium was refreshed every 24 hours.

### Human tissues

Healthy human lung tissues were obtained from residual donor lungs not used for transplantation. Diseased lung specimens were collected from patients with clinically confirmed PH who underwent lung transplantation at the First Affiliated Hospital of Zhejiang University School of Medicine (China) between March 2021 and December 2025 (IRB-2026-0080). PH diagnosis was confirmed based on clinical evaluation and histopathological features, including vascular remodeling and the presence of plexiform lesions, which were validated by α-SMA-IHC, and CD31-IHC staining. Exclusion criteria were predefined and included major comorbidities such as liver failure, end-stage renal disease requiring dialysis, active malignancy, chemotherapy, pregnancy, and lack of informed consent. This study was approved by the Institutional Review Board (Approval No. IBMS2018028), and all participants provided written informed consent for sample collection. All procedures involving human tissue adhered to the principles outlined in the Declaration of Helsinki [44]. Lung specimens were processed for multiple downstream applications. Tissues were either: (1) Snap-frozen in liquid nitrogen and stored at -80°C for molecular analysis; (2) Fixed in 4% paraformaldehyde (PFA; Servicebio, Cat. G1101) for paraffin embedding and histological evaluation; (3) Processed for single-cell RNA sequencing (scRNA-seq) and other assays, as detailed in subsequent sections.

### Echocardiography and haemodynamic measurements

Hemodynamic measurements were performed using established protocols. Following anesthesia, a Millar catheter (Smith Medical) was inserted into the right ventricle via the right jugular vein to measure RVSP. Continuous pressure signals were recorded and analyzed using a physiological data acquisition system. To evaluate right ventricular hypertrophy index (RVHI), hearts were excised post-mortem, and the right ventricle (RV) was carefully separated, rinsed in phosphate-buffered saline (PBS), and weighed. The RVHI was calculated using the following formula: RVHI = (right ventricular weight/ (left ventricle + septum weight)) × 100%^37, 38^. The total pulmonary vascular resistance index (tPVRI) was calculated using the following formula: tPVRI = RVSP (mmHg)/CO, where CI is RVCO (mL/min) * 100/body weight (g). RVSP was determined by right heart catheterization, and right ventricular cardiac output (RVCO) was assessed via transthoracic echocardiography.

### Immunofluorescence staining

Cryosections of mouse and human lung tissues were prepared for immunofluorescence staining. Briefly, tissues were embedded in optimal cutting temperature compound (OCT; SAKURA, 4583) and sectioned at a thickness of 4 μm using a cryostat (Leica CM1950, Leica Microsystems). The sections were air-dried at 37°C and subsequently blocked with 10% donkey serum (Servicebio, Cat. G1217) for 1 hour at room temperature (RT) to minimize nonspecific binding. After blocking, sections were incubated overnight at 4°C with primary antibodies diluted in antibody diluent. The following day, sections were washed and incubated with Alexa Fluor-conjugated secondary antibodies for 1 hour at RT. Nuclei were counterstained with DAPI, and slides were mounted using antifade mounting medium (Biosharp, Cat. BL701A). For co-localization of CD93 with general capillary (gCaps) markers (CD34 and APLNR), signal amplification was performed using an HRP-conjugated goat anti-mouse or anti-rabbit secondary antibody, followed by tyramide signal amplification (TSA) using the TSA kit (Servicebio, Cat. G1236) according to the manufacturer’ s instructions. Fluorescence images were acquired using an Olympus FV3000 OSR confocal microscope and processed using appropriate image analysis software. To ensure unbiased sampling, regions of interest were selected randomly. For quantification, at least three random fields of view were analyzed per sample. Investigators performing the staining and image analysis were blinded to the experimental groups.

### Chromatin Immunoprecipitation (ChIP) Assay

The ChIP assay was used to verify the transcriptional factor SMAD2 binding with the APLN gene. ChIP assays were performed using the SimpleChIP® Enzymatic Chromatin IP Kit (Cell Signaling Technology, Cat. #9003). For each immunoprecipitation, approximately 4 X 10 cells should be performed. The cells were first fixed with 1% formaldehyde for 10 min to crosslink protein to DNA and neutralized with for 5 minutes. Centrifuge cells at 2,000g for 5 min at 4°C, and the supernatant was resuspended.

Resuspend cells in 1 ml 1X Buffer A + DTT + PIC and incubate on ice for 10 min to pellet nuclei by centrifugation at 2,000g for 5min. Then, the samples were mixed with Micrococcal Nuclesse to digest DNA to length of approximately 150-900bp. Cell lysates were sonicated for 3 sets of 20-secpulses at a power of 35W to break nuclear membrane, and then incubated with SMAD2 antibody (Cell Signaling Technology, Cat. #5339) at a dilution of 1:50 overnight, followed by incubation with CHIP-Grade protein G Magnetic Beads. Normal Rabbit IgG (Cell Signaling Technology, Cat. #2729) was used as the negative control. The bound DNA-protein mixtures were eluted, and cross-linking was reversed after several washes. The purified DNA fragments obtained by ChIP were performed PCR and q-PCR analysis with APLN primers (5′ to 3′ sequences: TTTATGTGTGTGTGTGTGGAGGGG/TTCCAAGTGTTCAGCCTCCTCTTG). The PCR products were separated on 1 % agarose gels and visualized on a UV transilluminator.

To study whether SMAD2 can regulate the expression of APLN directly, we found that the APLN promoter region contains the SMAD2 binding site sequence (AGAC) by searching JASPAR (http://jaspar.genereg.net) (Fig. 6, E). A chromatin immunoprecipitation-PCR assay revealed that the biding of SMAD2 to the APLN promoter region was obvious versus IgG (Fig. 6, F and G)

### Statistical analysis

Statistical analysis were performed in GraphPad Prism 9. All experiments were performed triplicate and repeated at least 2 times. Normally distributed data, as determined by Shapiro-Wik test, were analyzed statistically using unpaired 2-tailed t tests. Non-normally distributed data were analyzed by 2-tailed Mann-Whitney U test. Multiple group analysis was performed by one-way ANOVA without post hoc correction for multiple comparisons. Data in figures are shown as mean ± SEM, as indicated. The specific statistical tests used are listed in the fiqure legends. Experiments were evaluated by statistical significance; P values less than 0.05 were considered significant.

## Data Availability

All data produced in the present study are available upon reasonable request to the authors

https://www.ncbi.nlm.nih.gov/geo/query/acc.cgi?acc=GSE169471

## Acknowledgments

We thank Zhongye Cai and Jingyu Chen for kindly providing PH patient samples. We thank Yan Zhang for providing the experimental drugs. We thank Xianbao Liu for providing supervision. We thank Robert Lafyatis for provide scRNA sequencing data. We thank Yingying Huang and Junli Xuan from the core facilities, Zhejiang University School of Medicine for their technical support.

This work was supported by the National Nature Science Foundation of China (82470431 to JY and 81670054 to JY), the National Key Research and Development Program of China (2023YFC2507100 to JYC, 2023YFC2507103 to JY, 2021YFA1100500 to XX and JY)

## Author contributions

JY conceptualized and designed the study, revised and finalized the manuscript, and provided financial support. YNL designed and performed the experiments, analyzed the data, and wrote the draft of the manuscript. XML helped with performing the mouse studies, cell culture, and tissue clearing 3D imaging. T.F.Z. helped with the analysis of scRNA transcriptome data. PM performed and analyzed the experiments related to hPSC. GYX helped with performing the mouse studies and cell differentiation experiment. XFF helped on animal catheterization. All authors contributed to manuscript revision.

## References

1 S. Rafii, J. M. Butler, and B.-S. Ding, Angiocrine functions of organ-specific endothelial cells. Nature. 529, 316–325 (2016)

2 J. C. Schupp, T. S. Adams, C. Cosme, M. S. B. Raredon, Y. Yuan, N. Omote, S. Poli, M. Chioccioli, K.-A. Rose, E. P. Manning, M. Sauler, G. DeIuliis, F. Ahangari, N. Neumark, A. C. Habermann, A. J. Gutierrez, L. T. Bui, R. Lafyatis, R. W. Pierce, K. B. Meyer, M. C. Nawijn, S. A. Teichmann, N. E. Banovich, J. A. Kropski, L. E. Niklason, D. Pe’er, X. Yan, R. J. Homer, I. O. Rosas, and N. Kaminski, Integrated Single-Cell Atlas of Endothelial Cells of the Human Lung. Circulation. 144, 286–302 (2021)

3 A. Mocumbi, M. Humbert, A. Saxena, Z.-C. Jing, K. Sliwa, F. Thienemann, S. L. Archer, and S. Stewart, Pulmonary hypertension. Nature Reviews Disease Primers. 10, (2024)

4 I. Borek, A. Birnhuber, N. F. Voelkel, L. M. Marsh, and G. Kwapiszewska, The vascular perspective on acute and chronic lung disease. Journal of Clinical Investigation. 133, (2023)

5 V. C. Ganta, W. S. Jones, and B. H. Annex, A conundrum of arterialized capillaries and vascular dilation in chronic limb-threatening ischaemia. European Heart Journal. 45, 265–267 (2024)

6 C. N. Hall, C. Reynell, B. Gesslein, N. B. Hamilton, A. Mishra, B. A. Sutherland, F. M. O’Farrell, A. M. Buchan, M. Lauritzen, and D. Attwell, Capillary pericytes regulate cerebral blood flow in health and disease. Nature. 508, 55–60 (2014)

7 Q. Zhang, N. Yaoita, A. Tabuchi, S. Liu, S.-H. Chen, Q. Li, N. Hegemann, C. Li, J. Rodor, S. Timm, H. Laban, T. Finkel, T. Stevens, D. F. Alvarez, L. Erfinanda, M. de Perrot, M. M. Kucherenko, C. Knosalla, M. Ochs, S. Dimmeler, T. Korff, S. Verma, A. H. Baker, and W. M. Kuebler, Endothelial Heterogeneity in the Response to Autophagy Drives Small Vessel Muscularization in Pulmonary Hypertension. Circulation. 150, 466–487 (2024)

8 A. L. Kern, D.-H. Park, J. Fuge, J. M. Hohlfeld, F. Wacker, M. M. Hoeper, K. M. Olsson, and J. Vogel-Claussen, Loss of pulmonary capillaries in idiopathic pulmonary arterial hypertension with low diffusion capacity is accompanied by early diffuse emphysema detected by 129Xe MRI. European Radiology. 35, 3010–3020 (2024)

9 S. Paolillo, S. Farina, M. Bussotti, A. Iorio, P. P. Filardi, M. F. Piepoli, and P. Agostoni, Exercise testing in the clinical management of patients affected by pulmonary arterial hypertension. European Journal of Preventive Cardiology. 19, 960–971 (2011)

10 A. Gillich, F. Zhang, C. G. Farmer, K. J. Travaglini, S. Y. Tan, M. Gu, B. Zhou, J. A. Feinstein, M. A. Krasnow, and R. J. Metzger, Capillary cell-type specialization in the alveolus. Nature. 586, 785–789 (2020)

11 J. Rodor, S. H. Chen, J. P. Scanlon, J. P. Monteiro, A. Caudrillier, S. Sweta, K. R. Stewart, A. Shmakova, R. Dobie, B. E. P. Henderson, K. Stewart, P. W. F. Hadoke, M. Southwood, S. D. Moore, P. D. Upton, N. W. Morrell, Z. Li, S. Y. Chan, A. Handen, R. Lafyatis, L. P. M. H. de Rooij, N. C. Henderson, P. Carmeliet, A. M. Spiroski, M. Brittan, and A. H. Baker, Single-cell RNA sequencing profiling of mouse endothelial cells in response to pulmonary arterial hypertension. Cardiovascular Research. 118, 2519–2534 (2022)

12 N. D. Cober, E. McCourt, R. S. Godoy, Y. Deng, K. Schlosser, A. Situ, D. P. Cook, S.-E. Lemay, T. Klouda, K. Yuan, S. Bonnet, and D. J. Stewart, Emergence of disease-specific endothelial and stromal cell populations responsible for arterial remodeling during development of pulmonary arterial hypertension. bioRxiv. 1–49 (2023)

13 F. Hua, S. Shang, Y.-w. Yang, H.-z. Zhang, T.-l. Xu, J.-j. Yu, D.-d. Zhou, B. Cui, K. Li, X.-x. Lv, X.-w. Zhang, S.-s. Liu, J.-m. Yu, F. Wang, C. Zhang, B. Huang, and Z.-W. Hu, TRIB3 Interacts With β-Catenin and TCF4 to Increase Stem Cell Features of Colorectal Cancer Stem Cells and Tumorigenesis. Gastroenterology. 156, 708–721.e15 (2019)

14 A. Petrović, J. Ban, M. Ivaničić, I. Tomljanović, and M. Mladinic, The Role of ATF3 in Neuronal Differentiation and Development of Neuronal Networks in Opossum Postnatal Cortical Cultures. International Journal of Molecular Sciences. 23, (2022)

15 K. Takahashi, and S. Yamanaka, A decade of transcription factor-mediated reprogramming to pluripotency. Nature Reviews Molecular Cell Biology. 17, 183–193 (2016)

16 B. Liu, D. Yi, X. Xia, K. Ramirez, H. Zhao, Y. Cao, A. Tripathi, R. Dong, A. Gao, H. Ding, S. Qiu, V. V. Kalinichenko, Y.-Y. Zhao, M. B. Fallon, and Z. Dai, General Capillary Endothelial Cells Undergo Reprogramming Into Arterial Endothelial Cells in Pulmonary Hypertension Through HIF-2α/Notch4 Pathway. Circulation. 150, 414–417 (2024)

17 L. Brash, G. D. Barnes, M. J. Brewis, A. C. Church, S. J. Gibbs, L. S. G. E. Howard, G. Jayasekera, M. K. Johnson, N. McGlinchey, J. Onorato, J. Simpson, C. Stirrat, S. Thomson, G. Watson, M. R. Wilkins, C. Xu, D. J. Welsh, D. E. Newby, and A. J. Peacock, Short-Term Hemodynamic Effects of Apelin in Patients With Pulmonary Arterial Hypertension. JACC: Basic to Translational Science. 3, 176–186 (2018)

18 A. G. Japp, N. L. Cruden, D. A. B. Amer, V. K. Y. Li, E. B. Goudie, N. R. Johnston, S. Sharma, I. Neilson, D. J. Webb, I. L. Megson, A. D. Flapan, and D. E. Newby, Vascular Effects of Apelin In Vivo in Man. Journal of the American College of Cardiology. 52, 908–913 (2008)

19 T. L. Williams, D. Nyimanu, R. E. Kuc, R. Foster, R. C. Glen, J. J. Maguire, and A. P. Davenport, The biased apelin receptor agonist, MM07, reverses Sugen/hypoxia-induced pulmonary arterial hypertension as effectively as the endothelin antagonist macitentan. Frontiers in Pharmacology. 15, (2024)

20 H. Zhao, M. M. Chakinala, M. B. Fallon, S. M. Lin, J. S. Lee, and Z. Dai, Endothelial Heterogeneity in Pulmonary Hypertension. Arteriosclerosis, Thrombosis, and Vascular Biology. 46, 3–16 (2026)

21 J. D. W. Evans, B. Girerd, D. Montani, X.-J. Wang, N. Galiè, E. D. Austin, G. Elliott, K. Asano, E. Grünig, Y. Yan, Z.-C. Jing, A. Manes, M. Palazzini, L. A. Wheeler, I. Nakayama, T. Satoh, C. Eichstaedt, K. Hinderhofer, M. Wolf, E. B. Rosenzweig, W. K. Chung, F. Soubrier, G. Simonneau, O. Sitbon, S. Gräf, S. Kaptoge, E. Di Angelantonio, M. Humbert, and N. W. Morrell, BMPR2 mutations and survival in pulmonary arterial hypertension: an individual participant data meta-analysis. The Lancet Respiratory Medicine. 4, 129–137 (2016)

22 G. Wang, B. Wen, Z. Deng, Y. Zhang, O. A. Kolesnichenko, V. Ustiyan, A. Pradhan, T. V. Kalin, and V. V. Kalinichenko, Endothelial progenitor cells stimulate neonatal lung angiogenesis through FOXF1-mediated activation of BMP9/ACVRL1 signaling. Nature Communications. 13, (2022)

23 R. Zhang, Y. Shen, X. Zhou, J. Li, H. Zhao, Z. Zhang, J. Zhao, H. Jin, S. Guo, H. Ding, G. Nie, Z. Zhang, Y. Wang, X. Yan, and K. Fan, Hypoxia-tropic delivery of nanozymes targeting transferrin receptor 1 for nasopharyngeal carcinoma radiotherapy sensitization. Nature Communications. 16, (2025)

24 A. Harada, Y. Yasumizu, T. Harada, K. Fumoto, A. Sato, N. Maehara, R. Sada, S. Matsumoto, T. Nishina, K. Takeda, E. Morii, H. Kayama, and A. Kikuchi, Hypoxia-induced Wnt5a-secreting fibroblasts promote colon cancer progression. Nature Communications. 16, (2025)

25 T. Heimbucher, J. Hog, P. Gupta, and C. T. Murphy, PQM-1 controls hypoxic survival via regulation of lipid metabolism. Nature Communications. 11, (2020)

26 D. Saygin, T. Tabib, H. E. T. Bittar, E. Valenzi, J. Sembrat, S. Y. Chan, M. Rojas, and R. Lafyatis, Transcriptional profiling of lung cell populations in idiopathic pulmonary arterial hypertension. Pulmonary Circulation. 10, 1–15 (2020)

27 E. Langenkamp, L. Zhang, R. Lugano, H. Huang, T. E. A. Elhassan, M. Georganaki, W. Bazzar, J. Lööf, G. Trendelenburg, M. Essand, F. Pontén, A. Smits, and A. Dimberg, Elevated Expression of the C-Type Lectin CD93 in the Glioblastoma Vasculature Regulates Cytoskeletal Rearrangements That Enhance Vessel Function and Reduce Host Survival. Cancer Research. 75, 4504–4516 (2015)

28 S. Mumby, F. Perros, C. Hui, B. L. Xu, W. Xu, V. Elyasigomari, A. Hautefort, G. Manaud, M. Humbert, K. F. Chung, S. J. Wort, and I. M. Adcock, Extracellular matrix degradation pathways and fatty acid metabolism regulate distinct pulmonary vascular cell types in pulmonary arterial hypertension. Pulmonary Circulation. 11, 1–16 (2021)

29 Y. Sun, W. Chen, R. J. Torphy, S. Yao, G. Zhu, R. Lin, R. Lugano, E. N. Miller, Y. Fujiwara, L. Bian, L. Zheng, S. Anand, F. Gao, W. Zhang, S. E. Ferrara, A. E. Goodspeed, A. Dimberg, X.-J. Wang, B. H. Edil, C. C. Barnett, R. D. Schulick, L. Chen, and Y. Zhu, Blockade of the CD93 pathway normalizes tumor vasculature to facilitate drug delivery and immunotherapy. Science Translational Medicine. 13, (2021)

30 Y. Xu, Y. Jia, N. Wu, J. Wang, L. He, and D. Yang, CD93 Ameliorates Diabetic Wounds by Promoting Angiogenesis via the p38MAPK/MK2/HSP27 Axis. European Journal of Vascular and Endovascular Surgery. 66, 707–721 (2023)

31 S. Helleberg, A. Engel, S. Ahmed, A. Ahmed, and G. Rådegran, Higher plasma IL-6 and PTX3 are associated with worse survival in left heart failure with pulmonary hypertension. American Heart Journal Plus: Cardiology Research and Practice. 20, (2022)

32 R.-H. Xu, T. L. Sampsell-Barron, F. Gu, S. Root, R. M. Peck, G. Pan, J. Yu, J. Antosiewicz-Bourget, S. Tian, R. Stewart, and J. A. Thomson, NANOG Is a Direct Target of TGFβ/Activin-Mediated SMAD Signaling in Human ESCs. Cell Stem Cell. 3, 196–206 (2008)

33 X. Ni, Y. Xu, W. Wang, B. Kong, J. Ouyang, J. Chen, M. Yan, Y. Wu, Q. Chen, X. Wang, H. Li, X. Gao, H. Guo, L. Cui, Z. Chen, Y. Shi, R. Zhu, W. Li, T. Shi, L.-F. Wang, J. Huang, C. Dong, and Y. Lai, IL-17D-induced inhibition of DDX5 expression in keratinocytes amplifies IL-36R-mediated skin inflammation. Nature Immunology. 23, 1577–1587 (2022)

34 W.-W. Wang, S.-Y. Ji, W. Zhang, J. Zhang, C. Cai, R. Hu, S.-K. Zang, L. Miao, H. Xu, L.-N. Chen, Z. Yang, J. Guo, J. Qin, D.-D. Shen, P. Liang, Y. Zhang, and Y. Zhang, Structure-based design of non-hypertrophic apelin receptor modulator. Cell. 187, 1460–1475.e20 (2024)

35 J. Huang, H.-y. Lee, X. Zhao, J. Han, Y. Su, Q. Sun, J. Shao, J. Ge, Y. Zhao, X. Bai, Y. He, X. Wang, X. Wang, and C. Dong, Interleukin-17D regulates group 3 innate lymphoid cell function through its receptor CD93. Immunity. 54, 673–686.e4 (2021)

36 Z. Hao, L. Ren, Z. Zhang, Z. Yang, S. Wu, G. Liu, B. Cheng, J. Wu, and J. Xia, A multifunctional neuromodulation platform utilizing Schwann cell-derived exosomes orchestrates bone microenvironment via immunomodulation, angiogenesis and osteogenesis. Bioactive Materials. 23, 206–222 (2023)

37 J. Zhang, P. Mao, T. Zhou, B. Yue, Y. Li, Y. Qiu, K. Ying, F. Wang, J. Chen, and J. Yang, Macrophage ferroptosis potentiates GCN2 deficiency induced pulmonary venous arterialization. Nature Communications. 16, (2025)

38 M. Du, H. Jiang, H. Liu, X. Zhao, Y. Zhou, F. Zhou, C. Piao, G. Xu, F. Ma, J. Wang, F. Perros, N. W. Morrell, H. Gu, and J. Yang, Single-cell RNA sequencing reveals thatBMPR2mutation regulates right ventricular functionvia IDgenes. European Respiratory Journal. 60, (2022)

